# Understanding Health worker Perspectives on Risk Screening for HIV Testing - A Qualitative Study from Zimbabwe

**DOI:** 10.1101/2022.05.09.22274852

**Authors:** Hamufare D. Mugauri, Joconiah Chirenda, Kudakwashe Takarinda, Owen Mugurungi, Getrude Ncube, Ishmael Chikondowa, Patrick Mantiziba, Mufuta Tshimanga

**Affiliations:** The University of Zimbabwe, Department of Primary Health-care Sciences, Harare, Zimbabwe; Ministry of Health and Child Care, AIDS and TB Unit, Harare, Zimbabwe; Organisation for Public Health Interventions and Development (OPHID), Harare, Zimbabwe; Clinton Health Access Initiative (CHAI), Harare, Zimbabwe

**Keywords:** Screening tool, qualitative, HIV Testing Services, health worker perspectives, grounded theory framework

## Abstract

**Background:** The use of screening tools during targeted human immunodeficiency virus (HIV) testing services improves efficiency by identifying individuals who are likely to test positive. Effective utilization of screening tools requires an understanding of health care worker perception and willingness to use the tools. We determined health workers’ perspectives on screening tools to enhance their effective and consistent utilisation.

**Methods:** We conducted a qualitative study among healthcare workers at 8 selected primary healthcare facilities in Zimbabwe. Interviewer guided, in-depth interviews were conducted with healthcare workers and their immediate supervisors. Inductive and deductive coding (hybrid) was applied to develop and analyze themes following a framework built around the grounded theory model to describe perspectives that influence the effective and consistent utilization of HIV screening tools and suggestions for improved eligibility screening.

**Results:** Behavioural factors facilitating the utilisation of the screening tool included motivation to adhere to standard practice, awareness of screening role in targeting testing, and its ability to manage workload through screening out ineligible clients. This was evident across service delivery levels. Barriers included limited healthcare capacity, confidentiality space, multiple screening tools and opaque screening in/out criteria and the potential of clients not responding to screening questions truthfully.

**Conclusions:** Across geographical and service delivery levels, correct placing of the screening tool at HIV testing entry point, healthcare worker knowledge on screening in/out criteria emerged as enablers for correct and consistent use of the screening tools. Further, standardizing the tools used would improve the utilisation of the correct tool.

## Introduction

More than two decades into the HIV pandemic, an estimated 79.3 million people have been infected and approximately 36.3 million people have succumbed to AIDS-related illnesses globally (1,2). The virus remains a major global public health threat with an estimated 37.7 million people living with HIV (including 1.7 million children), globally in 2020. Notably, around 16% of the people living with HIV (6.1 million) do not know their HIV status exposing a large gap in testing (1).

The HIV pandemic is skewed against East and Southern Africa, which contributes 20.6 million people living with HIV and 670,000 new HIV infections in 2020 alone, making it the epicentre of the pandemic (2). Further, one in every 25 adults (3.6%) is living with HIV in Southern Africa alone, accounting for more than two-thirds of the people living with HIV worldwide (1).

Knowing one’s HIV status through testing is key to mitigating the onward transmission of the virus in the community. While universal testing (provider and client-initiated testing) remains the gold standard, many resource-poor settings are struggling to offer this, mainly because of test kit shortages indicating the need for cost-effective approaches to HIV testing. To respond to this context, screening tools are suggested to aid testers to segregate clients and prioritize testing clients who are most likely to test HIV positive, thereby reducing “unnecessary testing” – whereby a negative test result is almost predictable. Screening tools are an integral component of the targeted testing strategy (3).

Zimbabwe shifted from testing for coverage and embraced targeted testing in 2017 as a stratagem to enhance positivity yield (4). Further, an adult HTS screening tool was introduced in 2019 to aid testers to direct HIV testing for clients likely to test positive. This tool was subsequently evaluated and validated, resulting in a revised tool that met the properties acceptable to effectively reduce testing volumes and minimally screen out potential positive testers.

During the evaluation and validation exercise, it was anticipated that the positivity yield would decline since no screening was being done (both screened in and screened out clients were tested) in contrast with before the exercise when the screening tool guided eligibility for testing. However, it was noted with concern that a positivity yield of 7.53% was documented during the evaluation comparable with 7.68%, which was documented at the same facilities a month before the evaluation exercise. This finding strongly suggested that either the tool was not being routinely utilized as expected, or that the tool was not effective in its determination of eligibility for testing.

Using a qualitative research approach and drawing on perspectives from Nurse managers and testers, this study sought to generate an in-depth understanding of the factors that influence the utilisation of screening tools at public health facilities in Zimbabwe. The goal was to inform the effective, routine and standardized implementation of screening tools to guide targeted testing.

## Materials and Methods

### Study design and theoretical framework

This cross-sectional qualitative study using in-depth interviews (IDIs) sought to understand and describe the factors that influence health workers’ and their managers’ perspectives on the utility of HIV Testing Services screening tools. We applied objectivist and constructivist attributes of the Grounded theory adapted to our context. This facilitated the application of the comparative methodology that provided systematic guidance for gathering, synthesizing, analysing, and conceptualizing qualitative data to understand health workers’ perspectives on the use of Screening tools in HIV testing (5). The adaptation of the Grounded theory is illustrated in **Fig 1**.

**Fig 1.**
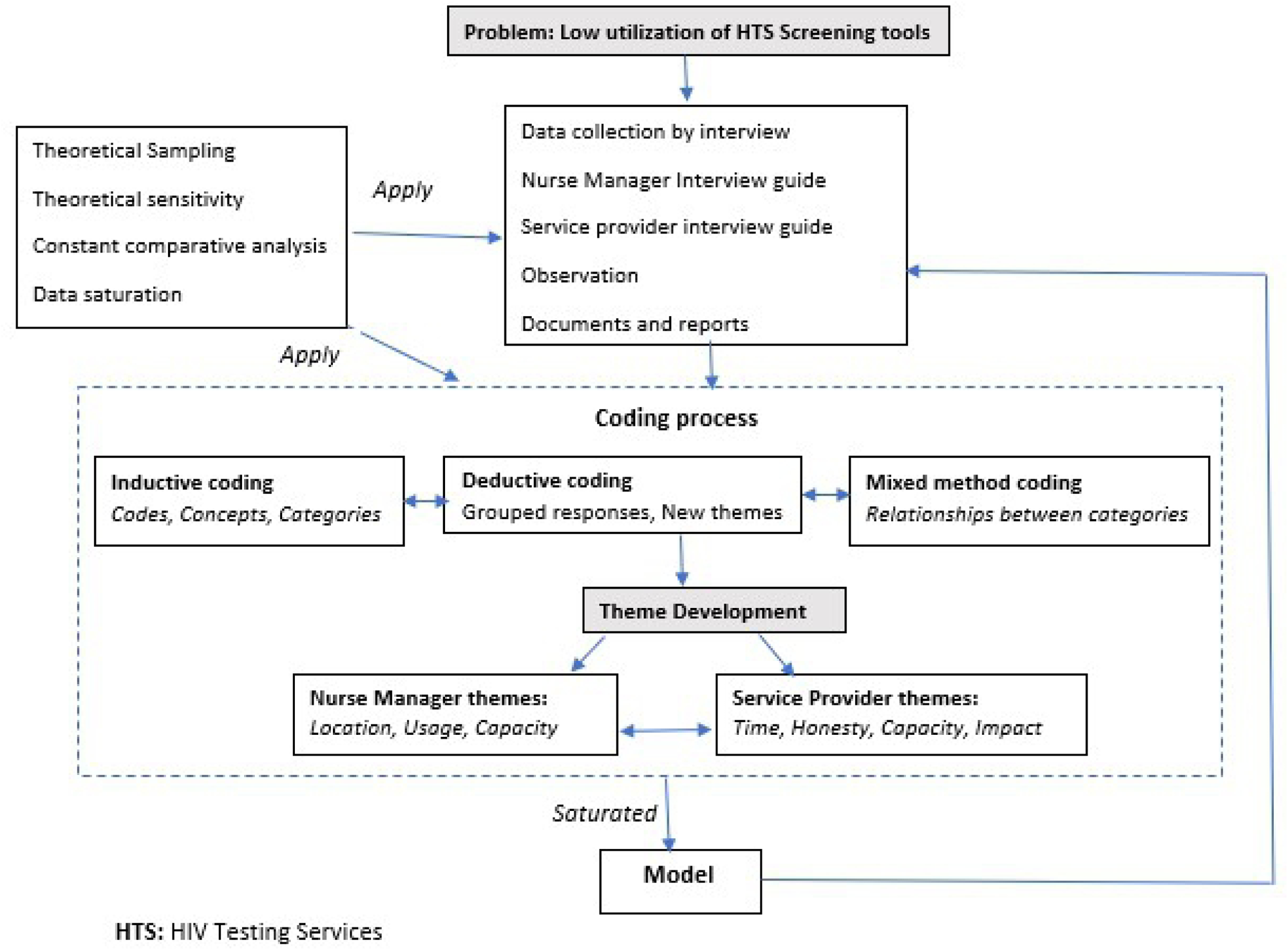
Adaptation of the grounded theory.

Two-part questionnaires were developed to guide the elicitation of key variables from the Nurse managers (Sister in Charge and Matrons) and the Testers (Nurses and Primary Counsellors).

### Study Setting

The study was carried out at Primary Health Care (PHC) facilities which are the first port of call for communities seeking healthcare in Zimbabwe.

Zimbabwe is a landlocked, low-income country in Southern Africa which is located between Botswana, South Africa, Mozambique and Zambia with an estimated population of 16 million and a human development index of 0.516, ranked number 154 globally out of 189 countries in 2016 (6). The country is divided into two urban provinces, eight rural provinces and 62 districts. The capital city is Harare and other major cities include Bulawayo, Gweru, Kadoma, Kwekwe, Masvingo and Mutare (7).

All clients who report at the public health facilities are offered HIV testing services after being screened for eligibility, according to existing Job aides and OSDM (Operational Service Delivery Manual) (8). Provider initiated testing and counselling (PITC) is practised at the facility and in the community, whereby the health worker makes the initiative to offer HIV testing services to eligible clients regardless of the purpose of the visit. Clients may also demand the service (Client-Initiated Testing and Counselling - CITC) (9). HIV screening results are not routinely documented, the process only aids the service provider to determine if the client can be tested during that visitor to be advised to report back at a later date, according to their risk profile.

Outpatients (OPD), Family and Child Health (FCH) departments, as well as Opportunistic Infections clinics (OIC), are the common entry points for HTS. Admitted clients may also be tested within the wards.

### Sampling, participant recruitment, and data collection

Eight healthcare facilities were selected from the 25 facilities that participated in the quantitative evaluation and validation of the screening tool. The rationale for facility selection was to synthesise interrelated circumstances and participants for the quantitative and qualitative studies, on account of their inter-relatedness. Health workers (Nurse managers and testers) found onsite during the data collection exercise were recruited into the study, which recorded a 100% response rate from the health workers identified. Data were collected in November 2021 by Data collectors with experience in conducting qualitative interviews. The facilities, selected from 4 of the 10 provinces of the country included 1 Rural hospital (Hwedza), 1 district Hospital (Banket), 1 Mission Hospital (Avilla), 3 urban Polyclinics (Zengeza, Overspill and Seke south), Partner run site (New Africa house Newstart centre) and a rural clinic (Ruyamuro) as tabulated below;

All participants were provided with detailed study information before giving their written informed consent. All participants were either Nurse managers (Sister in Charge or Matron) or Testers, (Nurses and Primary Counsellors) working at the selected clinics and willing to consent to the audio recording of the interview. Consenting participants were assigned a unique study number for confidentiality. The final sample of 20 participants included male and female Nurse managers, nurses and primary counsellors. The variedness of this ultimate sample would enable obtaining a fairly comprehensive picture of experiences and perceptions related to using HTS screening tools (10).

Interviews lasted 25–35 minutes and were carried out using a guide with open-ended questions. Topics covered in the guide included awareness of the existence of the screening tool, its usefulness and consistency in its usage to guide decision making on eligibility for an HIV test for clients. Experiences using screening tools as well as the barriers to and facilitators for usage, and provider perceptions of their value in targeting HIV testing were also investigated. Interviews were conducted in quiet locations, mostly in open spaces or in offices. Discussions were primarily conducted in English, but participants were free to express themselves in vernacular (Shona) which they felt helped them better articulate their experiences when utilising HTS screening tools. Saturation of themes during data collection was achieved through regular debriefing discussions among the investigators on probing techniques (11). Interviews were stopped when no new issues emerged.

### Data analysis

All interviews were transcribed verbatim. Audio recordings with renderings of local languages were directly transcribed and translated to English by the investigators fluent in the study languages, and checked the accuracy of the transcripts against digital recordings. Multiple reading of transcripts was done by both investigators, followed by manual coding and categorisation into pre-set themes, new themes were also developed from recurring related responses. Transcripts were imported into QSR International NVivo version 10 software to group the initial codes into themes and subsequently organize them into key dimensions and identify patterns across groups. (12). Soft-copy transcripts were stored securely and safely on password-protected computers and audio recordings were deleted from recorders. Transcripts were not returned to participants for comment.

The two members of the study team independently reviewed and coded the transcripts guided by the Grounded theory constructs to explore the perceptions of participants on perceptions on the utility of screening tools in public health settings. We applied open and axial coding to facilitate the interpretation of collected data. To analyse the qualitative data, we used thematic analysis and inductively and deductively developed codes (hybrid). The codes were organized into three overarching domains of factors for nurse managers, namely location, usage, and capacity. For implementers, the overarching themes were four, namely time, honesty, capacity and impact.

Collaboratively, the investigators reviewed and refined emerging key dimensions and themes The process of refining, and reviewing key dimensions and emerging themes were repeatedly done until saturation was achieved when no additional themes or categories could be identified (12). The analysis process identified salient differences in the health workers’ perceptions of screening tools and their utility in public health settings. Participant demographic characteristics were obtained from the qualitative interviews. We categorized gender based on the responses to the question: “Tell us more about yourself,” when the participant explicitly and voluntarily mentioned their gender as either male or female without probing.

## Results

### Participants’ characteristics

The participants (n = 20) were mostly female (n = 13, 65%), and of median age of 37 (IQR: 31–40) years. The majority were Primary Counsellors (n=9, 45%), followed by Sisters in Charge (n=6, 30%) and 1 Matron (5%). Most reported having professional experience of between 2 and 5 years (9, 45%) and having served for less than 2 years at the current facility (n=8, 40%). (Table 2.)

**Table 1.** Sites for Qualitative Data Collection for the Adult HTS Screening tool, 2021.

| Province | District | Name of site |
| --- | --- | --- |
| Mashonaland West | Banket | Banket District Hospital |
| Harare | Harare City | Ruyamuro clinic, Overspill Clinic, New Africa house<br>Newstart Centre |
|  | Chitungwiza | Seke South Clinic, Zengeza clinic |
| Manicaland | Nyanga | Avila Mission hospital |
| Mashonaland East | Hwedza | Hwedza Rural Hospital |

**Table 2.**
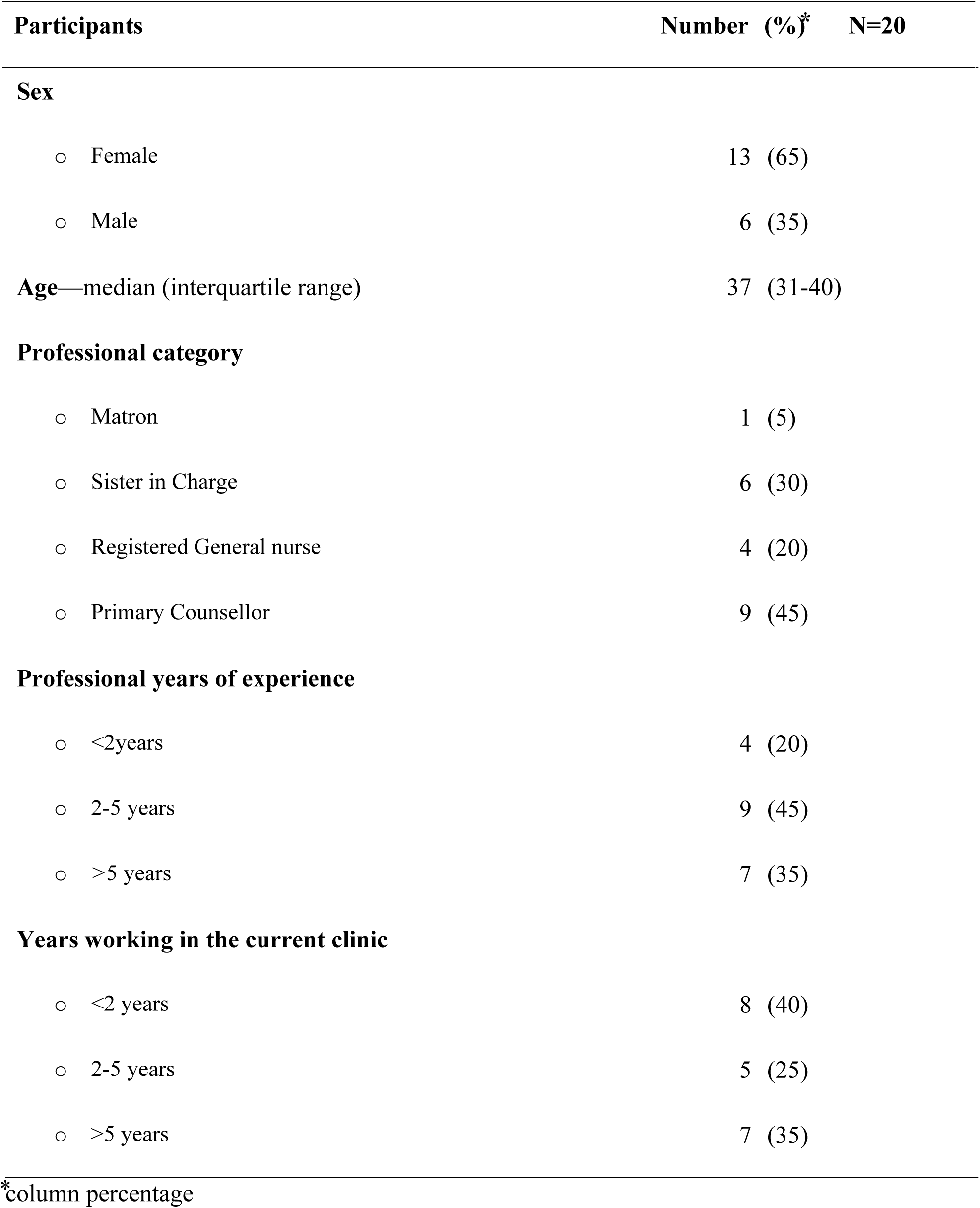
Demographic characteristics of participants.

We present five themes from the analysis (Table 3) supported with verbatim, minimally edited quotes.

**Table 3.** Themes and key dimensions from in-depth interviews and their relevant Grounded theory constructs and domains.

| Theme and key dimensions | Relevant Grounded theory construct and the *operational definition* | Relevant Grounded domain |
| --- | --- | --- |
| **Theme 1: Ideal placing of the HTS screening tool within the health care facility**<br>*Key dimensions*<br>Screening for HTS eligibility at facility entry, reception area, consultation room or HIV testing point<br>Opinions on the best placing of a screening tool within the healthcare setup | **Outcome expectations**<br>*“Health workers identifying the ideal location of the screening process to achieve optimal client flow and utility of the tool”* | Social or<br>Environmental<br>Factors |
| **Theme 2: Potential negative sequelae from utilising HIV screening tools by health workers**<br>*Key dimensions* | **Reciprocal determinism**<br>*“Interactions between personal and social/environmental factors that positively or negatively influence utilization of HIV screening tools”* |  |
| • Fear of the screening process increasing the workload for HIV testing<br>• Health workers are not clear about the screening in/out process due to a lack of orientation<br>• Concerns from providers that multiple tools are available and it's not clear which tool to utilize<br>• Client flow is already reduced at health facilities hence no need to screen the few that come |  |  |
| **Theme 3: Potential deliberate misinformation by clients desiring an HIV test**<br>*Key dimensions*<br>• Fear of clients not responding honestly when asked screening questions because of their desire to be tested/ not tested<br>• Confidentiality environment creation and assurance at the onset of engaging with the client<br>• Client attitudes towards being screened for eligibility before testing | **Behavioural capability**<br>*“Having and using acquired knowledge and skills to promote honesty in responding to screening questions to ensure that screening decision is based on true factors”* | Professional and Personal factors |
| **Theme 4: Amount of time required to perform the screening process** | **Self-efficacy** |  |
| *Key dimensions*<br>• To correctly ascertain the amount of time required to conduct HIV screening<br>• Contrasting the amount of time required to conduct an HIV test against the amount of screening<br>• Determining screening duration time reduction when screening is routinely performed | *“Having a good understanding of the importance of screening for HIV testing and the minimum time it takes when routinely applied”* |  |
| **Theme 5: The effect that screening for HIV testing has on various health aspects; resources, workload, efficiency**<br>*Key dimensions*<br>• Reflect on how reducing testing volumes through eligibility screening discourages high frequent testing with no corresponding positivity yield<br>• Drawing from regular onsite data analysis how positivity yield is impacted by testing volumes | **Observational learning**<br>*“Reflecting on the role of eligibility screening for HIV testing in reducing testing volumes, reduce workload and promote efficiency in HIV testing”*<br>**Reinforcements**<br>*“Encouraging positive changes through interpersonal and structural support”* | Environmental and professional factors |

### Health worker perceptions of screening tools

Healthcare Workers (HCW) expressed varying perceptions on the ideal placing of the screening tool to maintain an ideal client flow. Further, they expressed their opinions on the timeframe required to proficiently conduct the screening process and how the screening decision should be communicated to the client as well as how to deal with clients who may falsify responses to obtain the desired HIV test, or avoid it. The perceptions were informed by their experience using the tools and for others, how they perceived the questions when they were shared with them.

### The ideal placing of the HTS screening tool within the health care facility

This theme was observed across various facility levels and sizes. Clinic setups have single entry points and usually attend to low volume clients whereas larger facilities such as district and rural hospitals had multi-entry points hence the need to determine the most ideal placing of the tool.

> *“it is useful but needs to be placed at the right entry point, where the health worker engages with the client one on one…*.*” (Male, Primary Counsellor, District Hospital*.)

Further, a relationship between correct placing and subsequent utilization and the ease thereof was suggested.

> *“We need to screen clients at all testing points for HIV, where we meet the client who has opted in for HIV testing following the group education sessions. If we screen them on arrival, this may discourage them from visiting our facility. (Female, Sister in Charge, Rural Hospital)*

### Potential negative sequelae from utilising HIV screening tools by health workers

Participants across the geographical areas had different opinions on how screening tools would impact their workload which reflected that a few of them did not have a clear understanding of their utilization.

> *“When clients come into the testing room, they want to be tested. I would rather not waste time asking them screening questions when yet there is a queue outside…*.*” (Female, Primary Counsellor, Urban Clinic)*
>
> *“Few clients are turning up for HIV testing because of COVID-19. I think the few that come should just be tested because they made efforts to come. Those who are not at risk are not coming” (Male, Primary Care Nurse, Mission Hospital)*

Most of the health care workers were aware of the application of the screening tool to assess eligibility for an HIV test, which inevitably result in some clients being screened out.

> *“When a client is screened out, I won’t proceed with testing and explain that they are not eligible at the time”* – (*Female, Sister In Charge, Urban Clinic)*.

Few of the HCWs were not clear on the role of the screening tool: that it should be applied to assess eligibility for an HIV test on the day of the visit. If a client does not meet the screening criteria, then they should not be tested but advised on their next date for re-screening. Further, some clients should not be screened because they are catered for under specific programs.

> *“Client would still test despite being screened out according to SOP” Female, Sister In Charge, District Hospital)*.

The cited Standard Operating Procedure (SOP) referred to pregnant and lactating women retesting algorithm, which is unique to them whilst the rest of the population does not utilize it.

There seemed to be multiple screening tools being utilized, particularly among partner-run sites.

> *“Here we use our own tool supplied by our organisation, which is electronic because we review the work done by our counsellors in determining who to test and who not to test*.*”– (Female, Doctor, Partner-run health centre)*.

Lastly, discussions revealed that utilization of available interventions was said to depend on attitude and HIV risk perception.

### Potential deliberate misinformation by clients desiring an HIV test

Aligned to the behavioural capability construct, this theme focused on the risk of clients deliberately providing false information during the screening process to access an HIV test or decline it.

> “*Some clients will lie because they want to get tested and will be angry if you say you won’t test them” (Female, Registered General Nurse, Mission Hospital)*.

Health workers across geographical settings concurred that creating confidential space and assuring the client of the same is needed in routine practice when dealing with HIV issues and that the screening process is no exception.

> “*To get honest responses, we discuss with our clients in the privacy and assure them that no one will know about the conversation. We also explain that the risk assessment provides us with important information to advise them on how best to live their lives, without exposing themselves to HIV…” (Female Primary Care Nurse, Rural Hospital)*.

Discussions revealed that the screening process, just like any other medical procedure, requires confidentiality to be created and assured. Clients may vary their responses to achieve an end and it’s the health worker’s responsibility to identify inconsistencies and highlight them courteously to verify facts.

### Amount of time required to perform the screening process

Participants who had never used the screening tool were motivated to utilize the tool in pairs and determine how much time they required to apply it whilst those who had experience using the tool provided feedback on the time they usually took to complete the screening process.

> *“I only needed 6 minutes to ask all the questions because I was not familiar with them, with routine use, I will probably need less than 5 minutes because I would have memorised them…”(Male, Primary Counsellor, Rural Clinic)*.

Discussions revealed that health workers would take an average of 5 minutes if they routinely utilize the screening tool. Further, observing medical work ethics is essential to avoid the screening process being used to wantonly reduce workload.

> *“The time I need to conduct an HIV test is 25 minutes at the minimum, that is if I am doing things right, the screening time is less than a third of that time, so it’s not much, but there is a need to make sure everyone screened out was not eligible for a test, to avoid some people screening clients out to reduce workload” (Male, Primary Counsellor, Urban Clinic)*.

### The effect that screening for HIV testing has on various health aspects; resources, workload, efficiency

This theme focused on the impact of screening for eligibility for HIV testing on workload against positivity yielding results and efficiency in the delivery of HIV testing services. Consistency was observed, across geographical locations that screening and testing clients who are likely to test HIV positive result in efficiency and economic use of finite resources (test kits) whilst ensuring that the positivity yield is optimal.

> *“Seeing that our positivity remains low despite efforts to raise it, the screening will reduce the total number of tests we do and we will test clients who mostly test positive and we would have done well…”(Female, Matron, District Hospital)*.

HCWs mentioned additional strengthening of the existing system to ensure that screening becomes mandatory at all facilities and that the client responses to screening questions should be documented for verification.

## Discussion

Our findings highlight the relevance of using the GTM framework to enhance the routine utilization of HIV risk screening tools by HCWs. GTM provides a framework for understanding how perceptions about the ideal placing of the tool, self-efficacy and outcome expectations, influenced by personal, interpersonal and environmental factors, as well as behaviour capability, ultimately affect the utilization of the tools. In this regard, the FGDs revealed that placing the screening tool at the HIV testing entry points is ideal to ensure that the tool is administered to clients who are willing to conduct an HIV test and the screening process is conducted within a confidential space. Assuring the client of confidentiality was suggested to complement the environment and ensure that the client can freely discuss sexual matters. The relationship between confidentiality and client willingness to divulge sensitive information is well documented in the literature (13–15). The interactions between factors at each one of these levels are particularly important for understanding the factors that motivate the routine utilization of screening tools by HCWs in heterogeneous settings.

The construct of self-efficacy emphasized the importance of screening process orientation and awareness of the right tool for utilization. The existence of multiple screening tools was identified as a hindrance to the effective use of the same. Health workers across facility levels suggested the standardization of the screening tools across the country, regardless of whether a facility is supported by a partner or entirely run by the government. This will create a comprehensive database of screening for eligibility for testing thereby creating an opportunity to evaluate adherence to the laid down procedure at determined intervals.

The construct of behavioural capability emphasized the need for creating a therapeutic relationship with clients, grounded on confidentiality to ensure honesty in response to the screening questions. Inconsistencies in client responses to questions can be confronted in this confidential space complimented by assurance o confidentiality. This can only be achieved if the HCWs are skilled in counselling dynamics as emphasized in the literature (16,17).

Our study showed that the minimal time needed to conduct screening is +/-5mins. Routine implementation of the screening tool will result in the questions being integrated as part of a continuous therapeutic conversation with the healthcare worker, during which the risk profile of the client is determined and hence the screening decision arrived at. This finding was consistent with what is documented regarding the value of targeting HIV testing to high-risk clients who are likely to obtain a positive test result (18,19). Discussions with HCWs indicated that the time taken to screen is worth the benefits of screening out ineligible testers, improving efficiency in testing services and improving positivity yield since targeted testing is enhanced by testing individuals likely to obtain a positive diagnosis. Applied regularly and consistently, screening is an effective tool to improve client flow at health facilities.

Further, it was observed that the screening process needs to be integrated into the minimum package for clients seeking HIV testing services. To achieve this end screening should be a mandatory step for all clients seeking HIV testing services. This is consistent with the drive to target HIV testing where screening tools form an integral part of risk assessment, particularly in environments where clients have a culture of high-frequency testing without regard to risk. Done correctly and consistently, screening for eligibility for an HIV test has documented benefits (20,21).

## Conclusions

Assessing eligibility for an HIV test is an integral part of targeting HIV testing services. This reduces the retesting frequency and considers the risk profile before offering an HIV test. Across geographical and service delivery levels, the correct placing of the screening tool at HIV testing entry point, healthcare worker knowledge on screening in/out criteria emerged as enablers for correct and consistent use of the screening tools. Further, standardizing the tools used would improve the utilisation of the correct tool.

## Data Availability

All relevant data are within the manuscript and its Supporting Information files

## Declarations

### Ethics approval and consent to participate

Approval to conduct this study was obtained from the Ministry of Health and Child Care head office, the Joint Research Ethics Committee for the University of Zimbabwe Faculty of Medicine and Health Sciences and Parirenyatwa Group of Hospitals (JREC 280/2021) and the Medical Research Council of Zimbabwe (MRCZ/A/2783). Written informed consent was obtained from all participants before conducting FDGs and recording the participants.

### Consent for publication

All authors consent to the publication of this work

### Availability of data and material

The interview guide for this study has been provided in **S1 Annex**

### Competing interests

The authors declare that they have no competing interests

### Funding

This project did not receive direct funding for its implementation. The study was conducted as part of routine service provision for the Zimbabwe Ministry of Health and Childcare HIV testing services.

### Authors’ contributions

Conception and design: HDM, JC, KT, OM, GN, PM, MT; development of data capture tools: HDM, KT, PM; data collection: HDM, KT, PM; data entry: HDM, PM; data analysis and interpretation: all authors; preparing the first draft of the manuscript: HDM, KT, OM, JC, MT; critical review and approval of final draft: all authors

## Acknowledgements

I acknowledge several individuals and institutions that made this study a success. Special gratitude goes to my academic supervisors, Professor M. Tshimanga, Dr J. Chirenda and Dr K. Takarinda, The Director of AIDS & TB Unit, Dr O. Mugurungi and the entire HTS team for their support and prodding during this study. Further, I thank the Clinton Health Access Initiative which provided financial support to meet travelling costs associated with the implementation of this study.

**Supplementary file**: SI_ Qualitative Study Dataset

## Notes

### Competing Interest Statement

The authors have declared no competing interest.

### Funding Statement

The authors received no specific funding for this work.

